# Rapid assessment of COVID-19 mortality risk with GASS classifiers

**DOI:** 10.1101/2021.12.07.21267425

**Authors:** Salvatore Greco, Alessandro Salatiello, Nicolò Fabbri, Fabrizio Riguzzi, Emanuele Locorotondo, Riccardo Spaggiari, Alfredo De Giorgi, Angelina Passaro

## Abstract

**BACKGROUND:** Risk prediction scores and classification models are fundamental tools to effectively triage incoming COVID-19 patients. However, current triaging methods often have poor predictive performance, are based on variables that are expensive to measure, and lead to decisions that are sometimes hard to interpret.

**OBJECTIVE:** We introduce two new classification methods that are able to predict COVID-19 mortality risk from the automatic analysis of routine clinical variables with high accuracy and interpretability. The classifiers, denominated SVM22-GASS and Clinical-GASS, leverage machine learning methods and clinical expertise, respectively.

**METHODS:** Both classifiers were developed using a derivation cohort of 499 patients and were validated with an independent validation cohort of 250 patients. The cohorts included COVID-19 positive patients admitted to two hospitals in the Italian Province of Ferrara between March 2020 and June 2020 (derivation cohort) and between September 2020 and March 2021 (validation cohort). The potential predictive variables analyzed in this study included demographic, anamnestic, and laboratory data, retrieved with the patients’ consents from their electronic health records.

The SVM22-GASS classifier is based on a Support Vector Machine model (SVM) with Radial Basis Function kernel (RBF). Importantly, the model uses only a subset of predictive variables that were automatically selected with the Least Absolute Shrinkage and Selection Operator (LASSO), while the RBF kernel is approximated with random feature expansions to reduce the computational requirements. The Clinical-GASS classifier is a threshold-based classifier that leverages the General Assessment of SARS-CoV-2 Severity (GASS) score: a highly interpretable COVID-19-specific clinical score that has been recently shown to be more effective at predicting the COVID-19 mortality risk than standard clinical scores.

**RESULTS:** The SVM22-GASS model was able to predict the mortality risk of the validation cohort with an Area Under the Receiver Operating Characteristic Curve (AUC) of 0.87 and an accuracy of 0.88 — performing on par with influential classification methods that exploit variables derived from expensive analyses such as medical imaging. Furthermore, variable importance analyses showed that the model relies primarily on eight variables for its predictions: White Blood Cell Count, Lymphocyte Count, Brain Natriuretic Peptide, Creatine Phosphokinase, Lactate Dehydrogenase, Fibrinogen, PaO2/FiO2 Ratio, and High-Sensitivity Troponin I.

Similarly, the Clinical-GASS classifier predicted the mortality risk of the validation cohort with an AUC of 0.77 and an accuracy of 0.78 — on par with other established and emerging machine-learning-based methods.

**CONCLUSIONS:** Our results demonstrate that it is possible to accurately predict the COVID-19 mortality risk using only routine clinical variables that can be readily collected in the very early stages of hospital admission. The classifiers have the potential to assist the clinicians in quickly identifying the patients’ mortality risk to optimally allocate both human and financial resources.

## Introduction

In late 2019, a new member of the coronavirus family, named Severe Acute Respiratory Syndrome CoronaVirus-2 (SARS-CoV-2), emerged in the Chinese province of Hubei[1] and rapidly spread worldwide, causing the first pandemic by a coronavirus. As of November 2021, the virus has caused more than 250 million cases of infection, and has led to over 5 million deaths in more than 200 countries [2] ^2^. COVID-19 (COronaVIrus Disease 19), the clinical manifestation of SARS-CoV-2 infection, is undoubtedly the most important global health concern this century has witnessed, and has been taking a heavy toll in terms of human, financial, and social resources. Moreover, the appearance of new SARS-CoV-2 strains and the lagging vaccination campaigns hinder the objective of reaching herd immunity, implying that such issues will likely persist in the coming months.

Hospitals are particularly affected, especially during periods of increased disease transmission, where the number of patients that simultaneously need treatment increases exponentially. Such waves of patients can quickly deplete hospital resources if not dealt with optimally. In these circumstances, it is fundamental to estimate the amount of resources incoming COVID-19 patients will likely require during their hospital stay, ideally, in the early phases of hospital admission. This ensures that high-risk patients will have access to adequate resources and that, in general, hospital resources are not misdirected. General risk scores and comorbidity indices, such as the Charlson Comorbidity Index (CCI)[3] ^3^, have been shown to be helpful in estimating the prognosis and thus the level of care COVID-19 patients require[4,5] ^4,5^. Similarly, pneumonia-specific risk scores such as the CURB-6[6] ^6^ and the CURB-65[7] ^7^ scores have been shown to have even greater prognostic ability[8,9] ^8,9^. Nevertheless, both kinds of risk scores tend to underperform clinical risk scores that are specifically targeted to COVID-19 patients[10–12] ^10-12^. This is in part due to the remarkable variability across patients of the SARS-CoV-2 manifestations.

The SARS-CoV-2 infection manifests itself heterogeneously: symptoms can range from being completely absent (up to 33% of total cases[13] ^13^) to being critical (up to 5% of total cases[14] ^14^), in which case they include respiratory failure, high fever, hyperinflammation, and multiorgan dysfunction. The percentage of critical cases increases substantially in the cohorts of hospitalized COVID-19 patients, reaching peaks close to 20%[15] ^15^; such percentages are particularly high in older patients with coexisting morbid conditions and cardiovascular diseases. To move beyond the limitations of general risk scores, in this work, we introduce two COVID-19-specific classifiers: the Clinical-GASS and the SVM22-GASS. Both classifiers identify high-risk patients by predicting their 30-day mortality outcome and were developed and validated using independent derivation and validation cohorts. The SVM22-GASS classifier is based on a Support Vector Machine model (SVM) and is built in a fully data-driven fashion exploiting state-of-the-art machine learning methods. The Clinical-GASS builds on a COVID-19 specific risk score we recently introduced and proved effective at stratifying the population of hospitalized COVID-19 patients: the General Assessment of SARS-CoV-2 Severity (GASS) score[10] ^10^.

## Materials and methods

### Study design and participant recruitment

This is a prospective observational cohort study, developed in the two hospitals of Ferrara’s territory dedicated to COVID inpatients, “Arcispedale S.Anna” in Cona (Fe) and “ Ospedale del Delta” in Lagosanto (Fe). The province of Ferrara is geographically located in the Eastern part of the Emilia-Romagna region of Italy, with a population of approximately 350,000 inhabitants, and it is characterized by a high presence of elderly subjects (∼26% of total population is aged >65 years, and nearly 1% >90 years).

This study analyzes the data of two independent cohorts of COVID-19 patients: a *derivation cohort* — used to develop the classifiers — and a *validation cohort* — used to validate them. In the machine learning community, such data are commonly referred to as training and test set, respectively.

The derivation cohort comprises data of 499 patients recruited between March 2020 and June 2020, which have been already partially described and analyzed in our previous study[10] ^10^. The validation cohort comprises data of 450 patients recruited between September 2020 and March 2021; however, the data relative to 200/450 patients were not complete of all the relevant variables and were thus discarded.

The collected variables include demographic, anamnestic and laboratory data. These were extracted with the patients consent from their electronic health records. Baseline symptoms and vital signs were added to their records and used to calculate both their CCI score (Charlson Comorbidity Index), and the GASS score. Information about the items considered by CCI and GASS scores can be found in the *Supplementary Table*.

SARS-CoV-2 infection was confirmed with the reverse transcriptase-polymerase chain reaction (RT-PCR) test. The exclusion criteria were the negativity of the swab tests to viral detection and age (patients younger than 18 years were excluded). All patients signed an informed consent when possible; in case the patients were unable to sign, the consent was obtained from their legal representatives.

### Descriptive analyses

We evaluated the differences between subjects in terms of the three major COVID-19 outcomes (1. IoC=intensification of care, meant as the need for non-invasive mechanical ventilation or for endotracheal intubation; 2. in-hospital death; 3. 30-day death). The observation period was protracted until the 30^th^ day since hospital admission for those patients who survived the hospitalization: this was possible using records of the local registry office linked to the hospital informatic system.

Patients needing IoC can be recognized by the acronym “IoCp” while, for those who did not undergo IoC, we chose the acronym “nIoCp”. The groups of patients who survived or died after a period of observation of 30 days are presented with the acronyms “30-ddp” (30-day deceased patients) and “30-dsp” (30-day survived patients), respectively.

### Statistical analysis

Data analyses were performed by using SPSS 26.0 software (IBM SPSS Statistics, IBM Corporation) and MATLAB (MATLAB 2020a, The MathWorks, Natick, MA).

The normal distribution of the continuous variables was analyzed using Kolmogorov-Smirnov and Shapiro-Wilk tests. Variables not normally distributed were log-transformed before entering the parametric statistical analysis. Categorical variables were summarized by using frequencies and percentages, and continuous data were presented as median (interquartile range, IQR). The Mann-Whitney U test was used for continuous variables, while the χ2 test was used for categorical variables. Variables with a *p* value <0.05 in the univariate analyses were used to perform multivariate logistic regression analyses. All *p* values <0.05 are considered statistically significant.

### The Clinical-GASS classifier

In our previous study[10] ^10^, we discussed the ability of the GASS score to stratify the population of hospitalized COVID-19 patients into groups with significantly different 30-day mortality risk. Such a stratification has the potential to improve patient care by signaling to the hospital personnel the patients who are likely to need a mild (GASS < 6), moderate (6 ≤ GASS ≤ 10) and high (GASS>10) level of care. However, clinical personnel might also benefit from knowing whether the patient at hand has a low or a high risk of dying, and is likely to require specialized treatment such as the admission to the intensive care unit. This can be accomplished by building a *binary classifier* that predicts, for example, the 30-day mortality outcome. This is the primary outcome measure we aim to predict with this and the other classifiers considered in the following sections.

To assess the potential of the GASS score to be used for such a binary classification task, we built a simple threshold-based classifier. The classifier — named Clinical-GASS — was designed by simply computing the optimal Receiver Operating Characteristic (ROC) threshold on the training set, in terms of the cost/benefit ratio[16] ^16^; equal costs were assumed for the misclassification of survivors and non-survivors. Patients with a GASS score below such a threshold are classified as likely survivors while patients with a GASS score above such a threshold are considered as likely non-survivors.

A similar approach was used to build the Charlson Comorbidity Index (CCI) classifier from the CCI score[3] ^3^. The CCI is a clinical score that is often used as a baseline method to estimate the mortality risk in the general population.

### The SVM22-GASS classifier

Both the Clinical-GASS and the CCI classifiers are built leveraging medical expertise and thus might have a strong appeal to clinicians: they are likely to have full understanding of how the classifiers work, to trust their decisions and to thus be willing to use them. However, both classifiers only use a limited number of features (Clinical-GASS: n=11, CCI classifier: n=19) and combine them with only sums and other simple operations defined by piecewise constant functions. For these reasons, neither classifier might be able to fully exploit useful features contained in the patients’ health records, and to capture the non-linear interactions between the multitude of variables potentially at play in determining a patient’s survival outcome. To address this issue, we designed a Support-Vector-Machine-based classifier (SVM) with Radial Basis Function (RBF) kernel[17] ^17^. An RBF-SVM is a binary non-linear classification method that classifies data by non-linearly mapping the feature vectors into an infinite-dimensional space in which data from different groups can be easily separated by a hyperplane.

In brief, data *x* are classified by the decision function *f*(*x*), which is given by

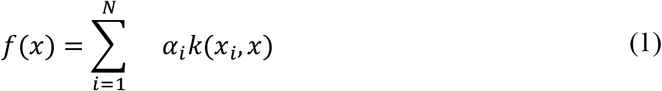

Here, *N* is the number of support vectors, *α*_*i*_ are weighting coefficients, and *k*(*y, z*) is the kernel function. In our case, we chose the Gaussian Radial Basis Function:

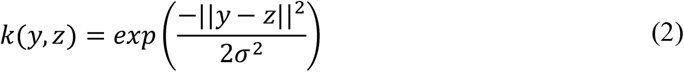

The SVM classifier was trained using only the data contained in the *training set*, was tested on the independent *test set*, and was compared against the CCI and GASS classifiers.

#### Data preprocessing

Before training the SVM classifier, we made sure to have sound training data. This was accomplished in two steps. First, we excluded from the analysis the variables that were measured in less than 50% of the patients. Secondly, we imputed the remaining missing values with the weighted average of the values of the three most similar instances, with weights inversely proportional to the distances from these. As a distance measure, we used the Euclidean distance for continuous features and the Hamming distance for binary ones.

#### Data augmentation

Learning a classifier from imbalanced training data such as ours (% survivors = 75.7 %) is a challenging task and can lead to poor sensitivity to the minority class. To deal with this issue, we balanced the training set using the Synthetic Minority Over-sampling Technique (SMOTE[18] ^18^). In brief, this method works by generating synthetic instances of the minority class by perturbing the minority instances in the directions of the difference vectors from their nearest neighbours of the same class.

#### Feature selection

To remove potentially redundant and uninformative features, while reducing the computational cost of training the SVM model, we performed feature selection; this was accomplished by learning a regularized logistic regression model with LASSO penalty[19,20] ^19,20^. The regularization parameter λ_LASSO_, which determines the regularization strength and thus the number of selected features, was chosen by minimizing the ten-fold cross-validation deviance.

#### Parameter training

To learn the model’s parameters, we minimized the hinge loss using the Limited-memory Broyden-Fletcher-Goldfarb-Shanno (LBFGS) solver[21] ^21^. Importantly, to speed up training and reduce memory requirements, we approximated the Gaussian kernel using the Fastfood[22] ^22^ random feature expansion method. To deal with potential residual classification biases due to the class imbalance, we adjusted the SVM classification threshold[23] ^23^. Specifically, we used the SVM classification scores to compute the optimal operating point of the ROC curve and used the resulting threshold to classify the data.

#### Hyperparameters optimization

Before learning the final model’s parameters, to preselect a class of SVM models suitable for the dataset at hand, we performed hyperparameter optimization[24] ^24^ using Bayesian optimization[25] ^25^. Specifically, with this procedure we found the kernel scale σ^2^, the regularization strength λ_SVM_, and the dimensionality of the random features space δ, that minimize the five-fold cross-validation hinge loss.

#### Model evaluation

To evaluate the quality of the model’s predictions, we computed Area Under the ROC Curve (AUC), accuracy, sensitivity, and specificity on the independent test set. The SVM’s performance is then compared to that of the other classifiers of interest, namely, the CCI classifier and the GASS classifier. As a baseline model, we also considered the majority classifier, which assigns the majority class to every instance in the dataset, regardless of its features.

#### Model interpretation

As it is often the case with most high-dimensional and non-linear machine learning models, also in our case, once the SVM model is fully trained, it is challenging to gain an intuitive understanding of the mechanism underlying the classifier’s decision process. To shed some light on such a mechanism, we estimated the variable importance (VI)[26] ^26^ of each input feature of the SVM model, by computing the *model reliance*. In brief, such method assigns high reliance to the features that, when perturbed. lead to a strong decrease in classification accuracy. Furthermore, to assess the discriminative power of the most important features selected with this method, we trained a reduced SVM model to classify survivors and non-survivors using only the top-3 features[27] ^27^ and measured the decrease in classification performance.

### Guidelines and Ethical approval

For the compilation of this manuscript, we followed STROBE (Strengthening the Reporting of Observational Studies in Epidemiology) guidelines for reporting observational studies.

The local Ethics Committee “Comitato Etico Indipendente di Area Vasta Emilia Centro (CE-AVEC)” approved the protocol of this study; the protocol code is 712/2020/Oss/AOUFe.

## Results

### Descriptive analyses

The distribution of males and females in the overall population was quite homogeneous (54.8% males vs. 45.2% females), while in the group of patients who underwent intensification of care (IoCp) there was a higher percentage of males (65.8% vs. 34.2%; *p*=0.021). The median age of the overall population was 72 years (IQR 58-82 years) and in-hospital mortality was recorded for 62 patients (24.8%)

The subjects who died within the hospitalization period (deceased) were significantly older than those who were discharged (median age 82 vs. 67 years; *p*<0.001), as well as it happened for the group of patients who died within 30 days (30-ddp) compared with those who survived within the same period (30-dsp) (median age 81 vs. 68 years; *p*<0.001).

Not surprisingly, we found differences between the IoCp and NIoCp groups in terms of length of stay (21 vs. 10 days; *p*<0.001); significant differences were also found between from the 30-ddp and 30-dsp groups (9 days vs. 13 days; *p*<0.001).

The GASS scores were higher in the IoCp group compared to NIoCp (9 vs. 7 points; p<0.001); as expected, similar differences were found between deceased and discharged patients (11 vs. 6 points; *p*<0.001) and also between 30-ddp and 30-dsp (11 vs. 7 points; *p*<0.001).

The patients with worse disease outcomes presented with a greater load of comorbidities (CCI 3.3±2.7 vs. 1.6±2.3 points, *p*<0.001 between deceased and discharged and 3.0±2.6 vs. 1.7±2.4 points, *p*=0.001 between 30-ddp and 30-dsp). As for vital parameters, IoCp and NIoCp groups differed in terms of respiratory rate, RR (22 bpm, IQR 20-30 vs. 22 bpm, IQR 16-22; *p*<0.001) and PaO2/FiO2 ratio at the first visit (300, IQR 230-352 vs. 229, IQR 155-295; *p*<0.001). Deceased and discharged patients differed in terms of systolic blood pressure (SBP), diastolic blood pressure (DBP) and RR; similarly, 30-ddp and 30-dsp differed for SBP, DBP and RR.

The laboratory findings were also heterogeneously distributed between groups: IoCp and NIoCp differed for serum C Reactive Protein (CRP), procalcitonin, D-Dimer, isoamylase, ALT, LDH and ferritin. Differences between deceased and discharged patients concerned serum lymphocyte count, creatinine, CRP, procalcitonin, D-Dimer, LDH, BNP and high sensitivity troponin I (HS TnI).

Similar differences were also found between 30-ddp and 30-dsp: these groups differed for serum creatinine, CRP, procalcitonin, D-Dimer (1.15 mg/L FEU, IQR 0.76-2.00 vs. 0.88 mg/L FEU, IQR 0.48-2.04; *p*=0.044), LDH, BNP and HS TnI (42 ng/ml, IQR 19-101 vs. 9 ng/ml, IQR 4-22; *p*<0.001).

All data concerning the characteristics of the studied population can be found in Table 1; extended results were reported only for those variables confirming their importance in the multivariate analyses later performed.

**Table 1.** Characteristics of population and differences between groups. Data are reported as median (interquartile range IQR) unless otherwise specified; p, p value between groups; IoCp, patients who underwent intensification of care; nIoCp, patients who did not undergo intensification of care; 30-ddp, patients deceased after 30 days; 30-dsp, patients survived after 30 days; GASS, General Assessment of SARS-CoV-2 patientS; SD, Standard Deviation; CCI, Charlson Comorbidity Index; SBP, Systolic Blood Pressure; DBP, Diastolic Blood Pressure; HR, Hart Rate; RR, Respiratory Rate; WBC, White Blood Cells; CRP, C Reactive Protein; ALT, Alanine Transferase; CPK, Creatine Phosphokinase; LDH, Lactic Dehydrogenase; BNP, Brain Natriuretic Peptide; HS TnI, High Sensitivity Troponin I.

| Cohort characteristics | All patients (N=250) | IoCp (N=76) | NIoCp (N=174) | *p* | Deceased (N=62) | Discharged (N=188) | *p* | 30-ddp (N=59) | 30-dsp (N=191) | *p* |
| --- | --- | --- | --- | --- | --- | --- | --- | --- | --- | --- |
| Males, n (%) | 137 (54.8) | 50 (65.8) | 87 (50.0) | 0.021 | 31 (50.0) | 106 (56.4) | 0.38 | 31 (52.5) | 109 (57.1) | 0.20 |
| Females, n (%) | 113 (45.2) | 26 (34.2) | 87 (50.0) |  | 31 (50.0) | 82 (43.6) |  | 28 (47.5) | 82 (42.9) |  |
| Age, years | 72 (58-82) | 72 (62-80) | 72 (53-82) | 0.26 | 82 (76-86) | 67 (53-78) | <0.001 | 81 (73-86) | 68 (54-79) | <0.001 |
| Length of stay, days | 12 (7-22) | 21 (12-40) | 10 (6-16) | <0.001 | 10 (6-20) | 12 (7-23) | 0.16 | 9 (6-16) | 13 (8-24) | <0.001 |
| GASS score, points | 7 (5-11) | 9 (7-11) | 7 (4-10) | <0.001 | 11 (9-13) | 6 (4-9) | <0.001 | 11 (8-13) | 7 (4-9) | <0.001 |
| CCI, points±SD | 2.0±2.5 | 2.1±2.5 | 2.0±2.5 | 0.69 | 3.3±2.7 | 1.6±2.3 | <0.001 | 3.0±2.6 | 1.7±2.4 | 0.001 |
| Vital Signs and Parameters |  |  |  |  |  |  |  |  |  |  |
| SBP, mmHg | 130 (120-140) | 130 (120-150) | 130 (115-140) | 0.47 | 120 (110-140) | 130 (120-140) | 0.024 | 120 (110-130) | 130 (120-140) | 0.005 |
| DBP, mmHg | 70 (70-80) | 70 (60-80) | 70 (70-80) | 0.08 | 70 (60-80) | 75 (70-80) | 0.019 | 70 (60-80) | 75 (70-80) | 0.003 |
| HR, bpm | 86 (77-99) | 86 (75-100) | 87 (78-99) | 0.39 | 88 (76-100) | 86 (78-99) | 0.94 | 88 (74-100) | 86 (78-99) | 0.84 |
| RR, apm | 20 (18-23) | 22 (20-30) | 20 (16-22) | <0.001 | 22 (18-28) | 20 (16-22) | <0.001 | 21 (18-28) | 20 (16-22) | 0.016 |
| PaO<sub>2</sub>/FiO<sub>2</sub> ratio | 300 (230-352) | 229 (155-295) | 310 (261-355) | <0.001 | 266 (200-304) | 310 (244-355) | 0.09 | 269 (193-311) | 308 (245-352) | 0.22 |
| Laboratory Findings |  |  |  |  |  |  |  |  |  |  |
| WBC (n/mm<sup>3</sup>) | 6395 (4815-9375) | 6970 (5315-9795) | 6210 (4750-9280) | 0.33 | 7600 (5380-10905) | 6140 (4750-9010) | 0.07 | 6280 (5060-10030) | 6420 (4795-9320) | 0.84 |
| Lymphocytes (n/mm<sup>3</sup>) | 1000 (740-1390) | 990 (635-1325) | 1000 (760-1470) | 0.33 | 870 (640-1085) | 1050 (778-1483) | 0.008 | 905 (658-1278) | 1030 (770-1465) | 0.14 |
| Creatinine (mg/dl) | 0.94 (0.77-1.28) | 0.96 (0.78-1.35) | 0.94 (0.77-1.24) | 0.78 | 1.34 (0.82-2.01) | 0.91 (0.75-1.16) | <0.001 | 1.31 (0.88-1.83) | 0.91 (0.75-1.17) | <0.001 |
| CRP (mg/dl) | 5.82 (1.98-10.92) | 8.57 (1.94-18.00) | 5.21 (1.98-9.52) | 0.038 | 8.49 (3.10-13.71) | 4.92 (1.74-9.87) | 0.007 | 8.49 (2.87-12.88) | 4.98 (1.83-10.12) | 0.025 |
| Procalcitonin (ng/ml) | 0.15 (0.05-0.51) | 0.25 (0.08-0.73) | 0.11 (0.05-0.43) | 0.018 | 0.54 (0.20-1.75) | 0.09 (0.05-0.26) | <0.001 | 0.49 (0.17-1.96) | 0.10 (0.05-0.27) | <0.001 |
| Fibrinogen (mg/dl) | 509 (441-659) | 555 (438-689) | 501 (441-643) | 0.27 | 572 (431-677) | 502 (441-642) | 0.37 | 550 (428-674) | 506 (441-643) | 0.91 |
| D-Dimer (mg/L FEU) | 0.93 (0.54-2.04) | 1.12 (0.74-1.99) | 0.88 (0.47-2.04) | 0.039 | 1.20 (0.77-2.30) | 0.85 (0.47-1.80) | 0.008 | 1.15 (0.76-2.00) | 0.88 (0.48-2.04) | 0.044 |
| Isoamylase (U/L) | 27 (20-44) | 36 (25-100) | 26 (18-41) | 0.008 | 32 (18-51) | 27 (21-44) | 0.92 | 36 (20-45) | 27 (20-43) | 0.63 |
| ALT (U/L) | 21 (14-37) | 26 (18-49) | 19 (12-33) | 0.003 | 18 (11-29) | 87 (52-193) | 0.06 | 19 (12-34) | 21 (14-37) | 0.29 |
| CPK (U/L) | 87 (51-203) | 111 (54-224) | 83 (46-169) | 0.14 | 93 (50-220) | 280 (211-361) | 0.82 | 122 (59-356) | 86 (50-167) | 0.12 |
| LDH (U/L) | 286 (219-380) | 390 (274-466) | 271 (210-332) | <0.001 | 328 (250-436) | 304 (125-593) | 0.015 | 332 (265-434) | 276 (210-358) | 0.001 |
| Ferritin (ng/ml) | 327 (142-637) | 506 (294-922) | 275 (120-565) | 0.006 | 364 (180-767) | 304 (125-593) | 0.19 | 439 (264-736) | 293 (121-610) | 0.07 |
| BNP (pg/ml) | 76 (36-185) | 80 (43-168) | 75 (33-195) | 0.90 | 197 (94-410) | 60 (30-126) | <0.001 | 209 (87-377) | 62 (30-132) | <0.001 |
| HS TnI (ng/ml) | 14 (5-34) | 15 (7-47) | 13 (5-29) | 0.13 | 41 (16-85) | 9 (4-21) | <0.001 | 42 (19-101) | 9 (4-22) | <0.001 |

Differences between groups were also found in terms of comorbidities, and all the results are summarized in Table 2: patients of the IoCp group were more often smoker; in the group of deceased subjects we found more diagnosis of hypertension, ischemic heart disease, heart failure, chronic kidney disease, history of stroke or transient ischemic attack (TIA), peripheral arterial disease (PAD), chronic obstructive pulmonary disease (COPD), localized or haematological cancer, dementia and diabetes with organ damage.

**Table 2.**
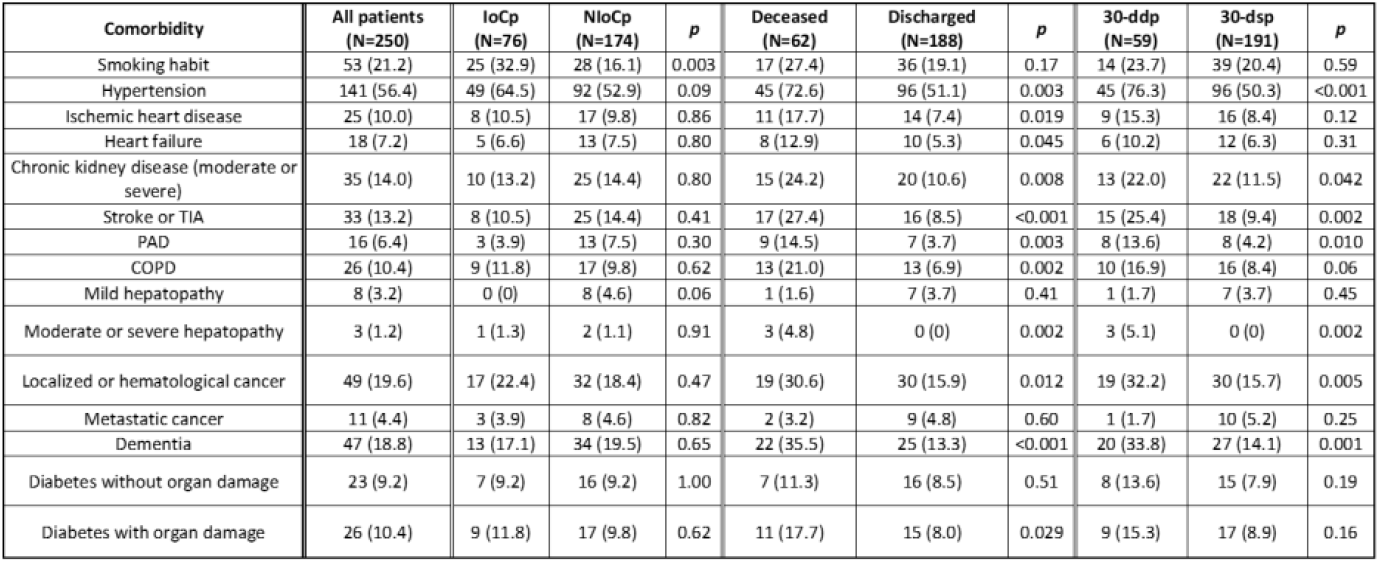
Comorbidities and differences between groups. Data are reported as number of subjects (percentage). p, p value between groups; IoCp, patients who underwent intensification of care; nIoCp, patients who did not undergo intensification of care; 30-ddp, patients deceased after 30 days; 30-dsp, patients survived after 30 days; TIA, Transient Ischemic Attack; PAD, Peripheral Arterial Disease; COPD, Chronic Obstructive Pulmonary DiseaseThe subjects from the 30-ddp group presented in their clinical history more often hypertension, chronic kidney disease, stroke/TIA, COPD, localized or haematological cancer and dementia.

Comparison analyses between groups were also performed for each item considered in the Clinical-GASS score and they can be found in the *Supplementary Table*. All variables found to be significantly different between groups in the univariate analyses were entered into multivariate analyses.

In the logistic regression analyses concerning the need for intensification of care, the strongest predictive variable was the PaO2/FiO2 ratio: both ratios <100 and 100-199 showed to be somehow protective towards the intensification of care (OR 0.11, 95% CI 0.02-0.72; *p*=0.021 and OR 0.07, 95% CI 0.02-0.27; *p*<0.001, respectively), while the male sex and a higher respiratory rate (>30 breaths per minute) seemed to be predictive for a higher need for intensification of care (OR 3.56, 95% CI 1.20-10.56; *p*=0.022 and OR 4.76, 95% CI 1.00-22.96; *p*=0.050).

Concerning the in-hospital mortality, age and PaO2/FiO2 ratio >300 were the only variables able to adequately predict this outcome (OR 1.07, 95% CI 1.02-1.12; *p*=0.007 and OR 0.35, 95% CI 0.13-0.92; *p*=0.034, respectively); moreover, age was particularly able at predicting death within a period of 30 days (OR 1.06, 95% CI 1.01-1.12; *p*=0.027), together with serum D-dimer between 0.5 and 2.0 mg/L FEU (OR 4.22, 95% CI 1.28-13.90; *p*=0.018); serum troponin I (HS TnI) < 20 ng/ml was, instead, protective towards 30-day death (OR 0.23, 95% CI 0.06-0.80; *p*=0.022). The forest plots concerning the regression analyses are reported in figure 1.

**Figure 1.**
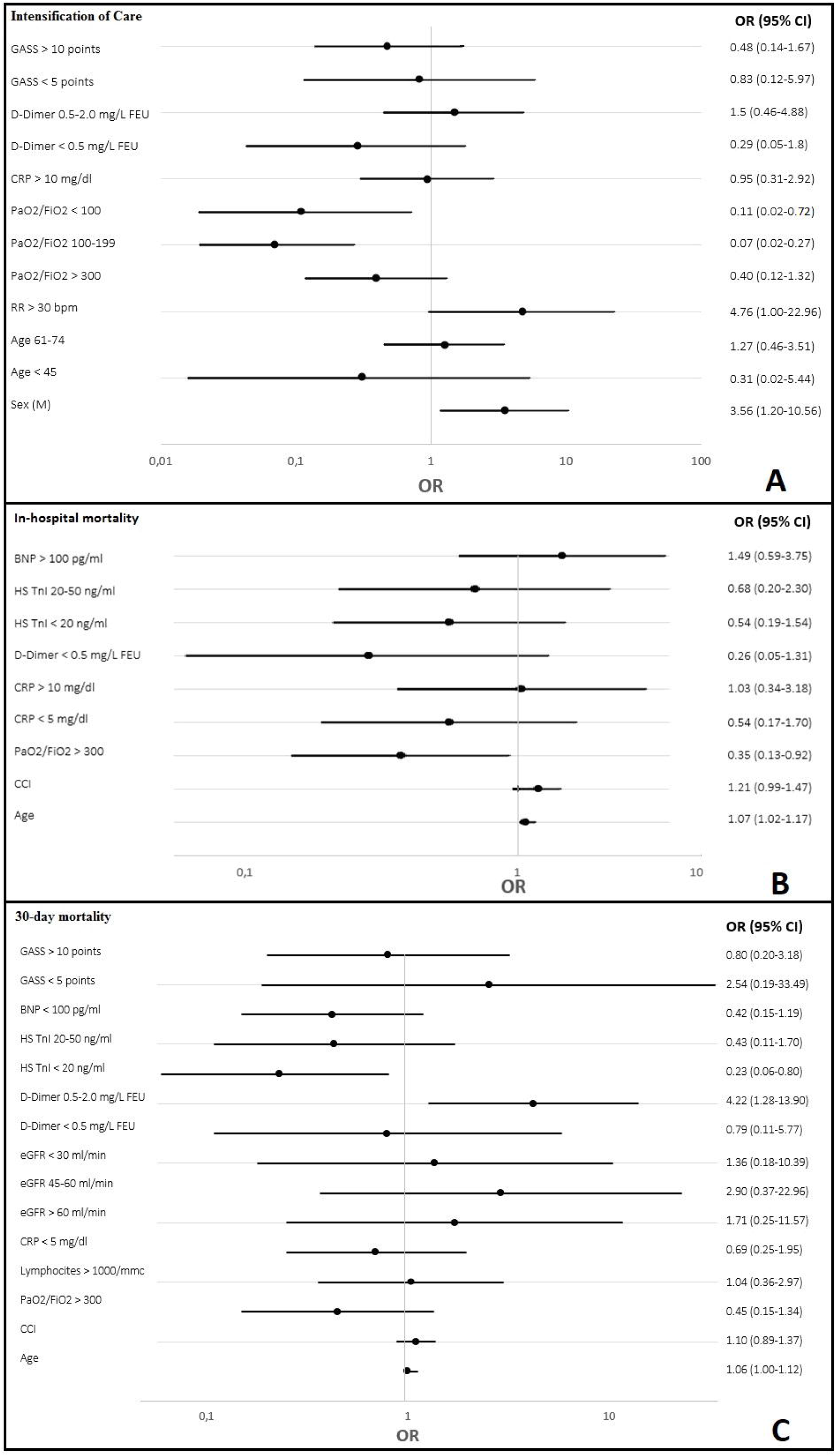
Forest plots of Regression Models. **(A)** Logistic regression modelling for identifying the variables associated with the need for intensification of care. **(B)** Logistic regression modelling for identifying the variables associated with in-hospital mortality. **(C)** Logistic regression modelling for identifying the variables associated with 30-day mortality.

The Clinical-GASS score was calculated for each patient, and this allowed us to predict the relative risk of in-hospital and 30-day death based on the analyses of this cohort of subjects. We developed an open access web tool in order to help clinicians identify the patients at higher risk of bad outcomes by COVID-19, simply filling out the form with all the variables needed. This tool can be found at the following link: https://ml.unife.it/GASS.html and it is available for free consultation.

Furthermore, we performed Receiver Operating Characteristic (ROC) curve analyses, in order to evaluate the predictive power of the GASS score, compared to the Charlson Comorbidity Index, which has been also recently tested in COVID-19 inpatients[28] ^28^. The main results of the classification analysis are reported in Figure 2.

**Figure 2.**
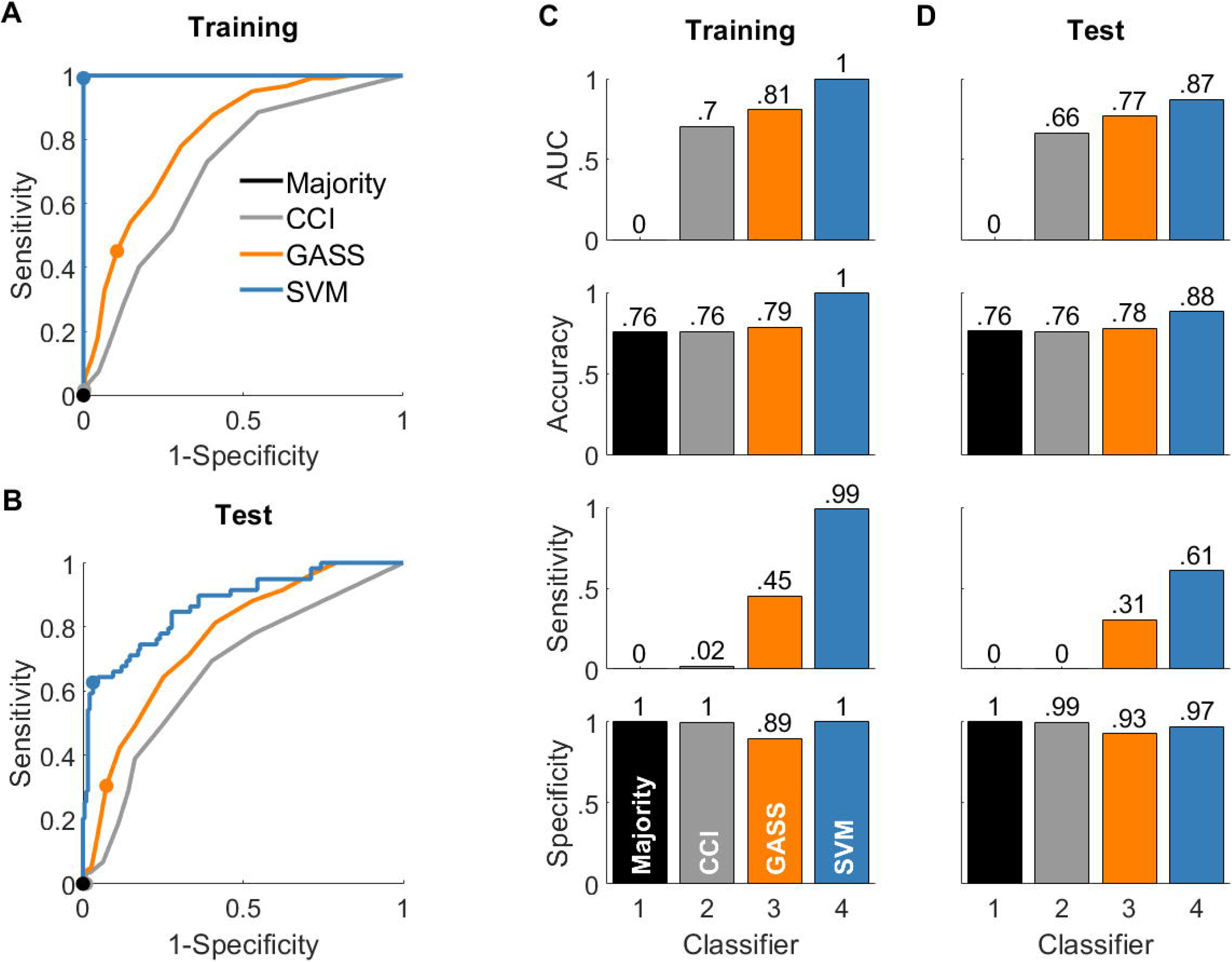
Summary prediction performance. **(A)** Receiver Operating Characteristic (ROC) curves of the considered classifiers, computed on the training set (majority classifier: black; Charlson Comorbidity Index (CCI) classifier: grey; GASS classifier: orange; SVM classifier: blue). Filled circles indicate the optimal operating points. Note that the classification threshold of the majority classifier is fixed; thus, the corresponding ROC curve collapses to a single point. **(B)** Corresponding curves computed on the independent test set. Filled circles represent the optimal operating points of the training ROC curves. **(C)** Considered classification performance measures, computed on the training set; values are rounded to the nearest hundredths; AUC: Area Under the ROC Curve. **(D)** Corresponding measures computed on the test set. Note that the majority class in both the training and test set is the survival class, which was arbitrarily associated to a negative test result; therefore, by always predicting survival, the majority classifier has a sensitivity of 0 and a specificity of 1.

### The CCI classifier

The optimal ROC threshold for the Charlson Comorbidity Index classifier was 11. This means that the optimal way to classify COVID-19 patients based on the CCI alone is to consider as high-risk all the patients with a CCI greater than or equal to this value. However, this classifier performed poorly on the test set, with an Area Under the Curve (AUC) of 0.66 and an accuracy of 0.76 (Figure 2D, grey bars). The poor performance of this classifier is attributable to the complete inability to correctly identify non-survivors (sensitivity=0), which makes it effectively equivalent to the naïve majority classifier (black bars).

### The Clinical-GASS classifier

The optimal ROC threshold for the Clinical-GASS classifier was 13, which lies, as expected, within the range of values of the high mortality risk class (GASS >10). Overall, the Clinical-GASS displayed satisfying test performance, with an AUC of 0.77 and an accuracy of 0.78 (Figure 2D, orange bars). Importantly, the Clinical-GASS classifier performed better than both the majority and the CCI classifiers, due to its improved ability to identify non-survivors (sensitivity=0.31).

### The SVM22-GASS classifier

The minimum-deviance ten-fold cross-validation regularization strength λ_LASSO_ was equal to 4.9·10^−3^. The corresponding LASSO logistic regression model fitted with such a parameter selected a total of 38 features, which are reported in Figure 2A. The best cross-validated hyperparameters of the RBF-SVM classifier trained using such features were σ^2^ = 774.16, λ_SVM_ = 1.4·10^−6^, and δ = 7318. The classifier trained with such hyperparameters achieved nearly optimal performance on the training set (AUC ≈ 1, accuracy ≈ 1, Figure 2C – blue bars), which means that the decision hyperplane is able to completely separate survivors from non-survivors in the transformed feature space. Importantly, the SVM classifier achieved very good performance also on the independent test set, with an AUC of 0.87 and an accuracy of 0.88. The ability to classify COVID-19 patients of the SVM classifier is thus superior to that of all the other considered classifiers. Such an improvement is largely attributable to a markedly better ability to correctly identify non-survivors (sensitivity=0.61).

#### Model interpretation and reduced models

The SVM classifier just described predicts the most likely 30-day mortality outcome of COVID-19 patients by non-linearly processing a selection of 38 features that characterize them. To understand whether some of these features are more important than others, we computed the model reliance[16]. The results of this analysis are reported in Figure 3A, where we plotted the model reliance of the full SVM model, on all the 38 features. The features are sorted from top to bottom in decreasing order of reliance. According to this analysis, 8 of the 38 features appear to be particularly informative to the model, as its predictions change drastically when such features are perturbed. Specifically, the eight most important features to the full SVM classifier are White Blood Cell Count (WBC), Lymphocyte Count (LYM), Brain Natriuretic Peptide (BNP), Creatine Phosphokinase (CPK), Lactate Dehydrogenase (LDH), Fibrinogen (FIBR), PaO2/FiO2 Ratio (PFR), and High-Sensitivity Troponin I (TnI).

**Figure 3.**
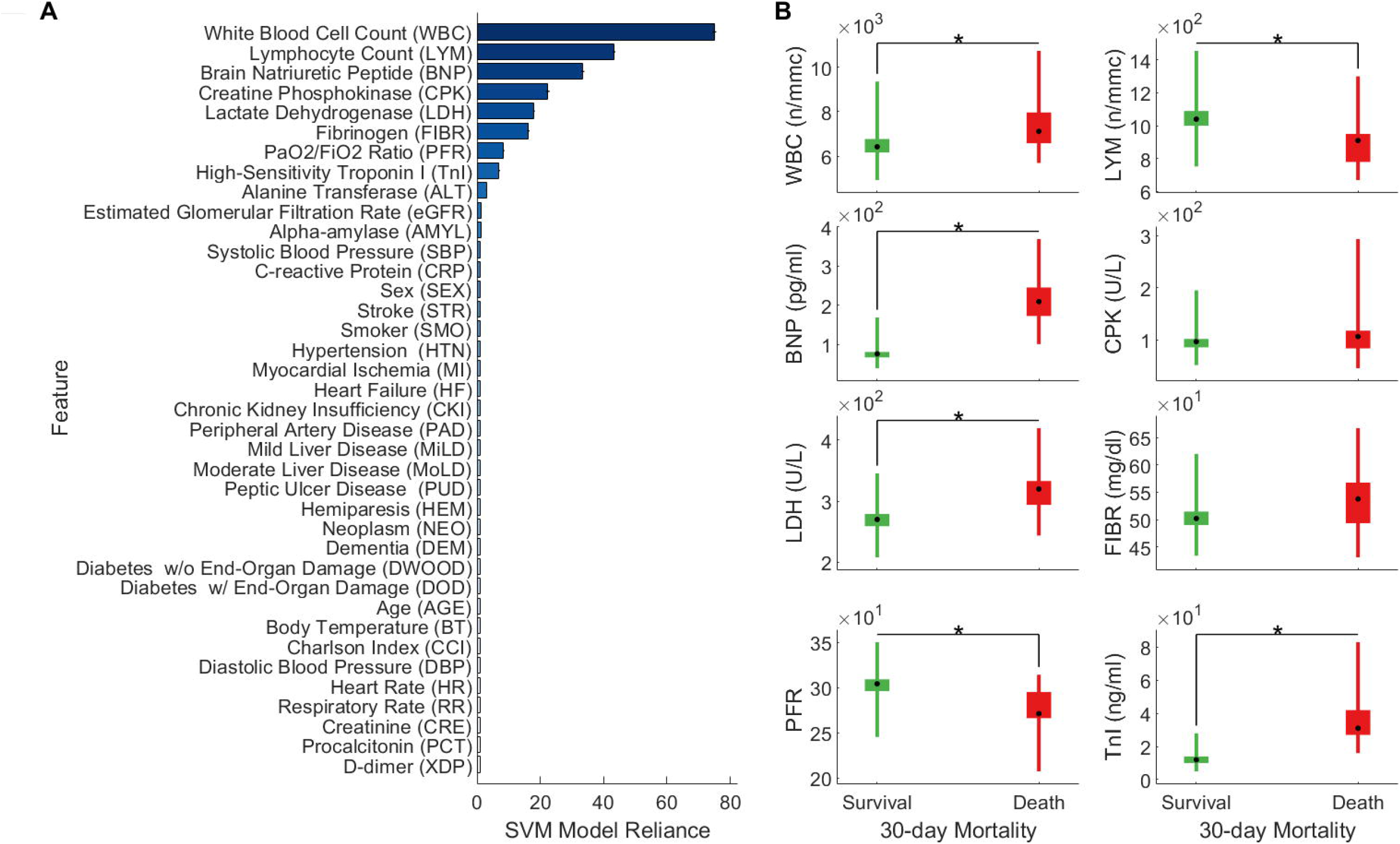
Understanding the full SVM model. **(A)** Model reliance of the full SVM model on all the 38 features its predictions are based on. Features are sorted (from top to bottom) in descending order of importance. **(B)** Medians, 95% bootstrap confidence intervals, and interquartile ranges of the top eight features, for the 30-day survival (green) and death (red) group. Black dots represent medians; boxes heights represent confidence intervals; thin lines represent interquartile ranges. Asterisks indicate significant (Bonferroni-adjusted *p* value ≤0.05) differences between groups (Mann–Whitney U test).

To understand how such features, differ between survivors and non-survivors, and whether such differences are statistically significant, we performed Mann–Whitney U tests, using a Bonferroni-adjusted significance value of 0.05 (Figure 2B). Out of the 8 most important features, only 2 do not appear to be significantly different between survivors and non-survivors (CPK and fibrinogen, p value >0.05). Medians, 95% bootstrap confidence intervals, and interquartile ranges of the top eight features, for the 30-day survival (green) and death (red) group can be found in the Figure 3, panel B.

The fact that the full SVM model appears to rely mostly on a subset of the full feature set of 38 features, raises the question of whether a reduced model, which has only access to the most informative features, can also accurately predict the COVID-19 outcome. To address this question, we trained a reduced RBF-SVM model to predict the COVID-19 outcome using only the three most informative features, that is White Blood Cell Count (WBC), Lymphocyte Count (LYM), and Brain Natriuretic Peptide (BNP). Choosing exactly three features allows us to visualize the entire feature space in which the classifier operates, and to plot the resulting decision surface.

As an additional baseline, we also trained another RBF-SVM model that had access to only the three least informative features [namely, Creatinine (CRE), Procalcitonin (PCT), and D-dimer (XDP)]. The models, which we refer to as SVM-TOP3 and SVM-BOT3 respectively, were trained following the same steps adopted for the full SVM model. The results of this analysis are reported in Figure 4. As expected, both reduced models exhibited worse training and test performance than the full SVM model (Figure 4B, 4C). Nevertheless, while the SVM-BOT3 classifier exhibited overall poor test performance, comparable to that of the CCI classifier (AUC = 0.7, accuracy = 0.7 — Figure 4C), the SVM-TOP3 classifier performed surprisingly well. As a matter of fact, the SVM-TOP3 had better test performance than the Clinical-GASS classifier (AUC = 0.83, accuracy = 0.83 — Figure 4C), despite using only three features.

**Figure 4.**
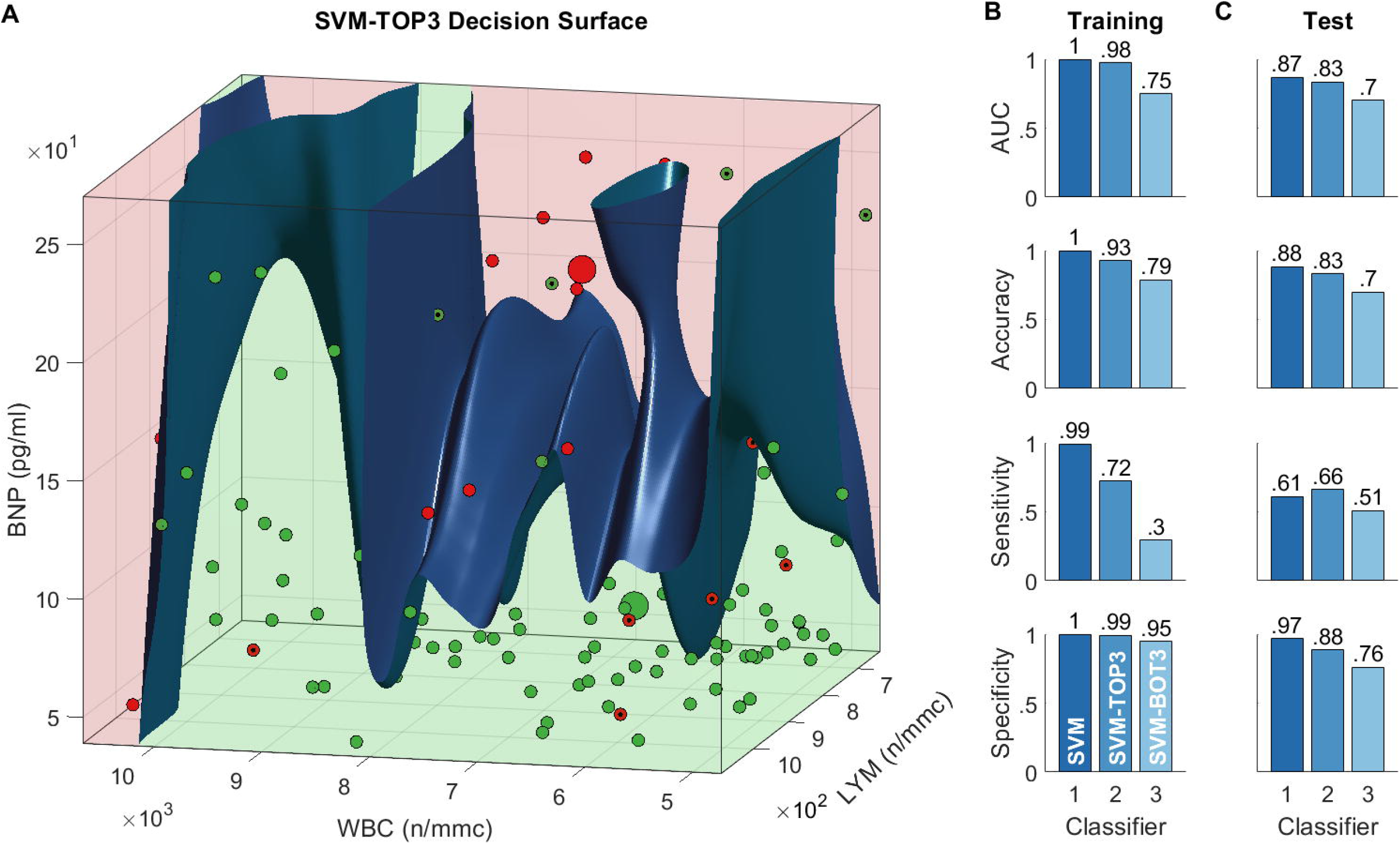
Reduced models and decision surface. **(A)** Decision surface of the SVM-TOP3 model, which only uses the top-3 features. The surface divides the feature space into two regions: a survival region (green) and a death region (red). Small green circles represent 30-day survivors, while small red circles represent 30-day non-survivors; misclassified instances are marked with a black dot. The corresponding large circles represent the group medians. Note: to facilitate the visualization of the decision surface, we are only plotting the data within a cuboid delimited by the 20^th^ and 80^th^ percentiles. **(B)** Classification performance measures of the full SVM model and of the reduced SVM models, computed on the training set. The SVM-TOP3 model only uses the top-3 features, while the SVM-BOT3 model only uses the bottom-3 features; values are rounded to the nearest hundredths. **(C)** Corresponding measures computed on the test set.

The resulting decision surface (Figure 4A) is, as expected, highly non-linear. The 3D plot of the feature space confirms that, generally, non-survivors seem to have higher serum BNP and lymphocyte count. Interestingly, the white blood cells count appears to become strongly predictive of a non-survival outcome, only for very high values (WBC>10·10^3^ n/mmc), especially when associated with high values of lymphocytes (LYM>9 · 10^2^ n/mmc).

## Discussion

Predicting the mortality risk of hospitalized COVID-19 patients could go a long way to optimally allocate hospital resources. Popular clinical scores are potentially useful in this regard, but have proven to be less accurate than COVID-19-specific methods[10–12] ^10-12^. In this work we have introduced and validated two new methods for predicting 30-day mortality risk: the Clinical-GASS and the SVM22-GASS. Overall, our results show that both methods are reliable and can thus be used to effectively triage incoming COVID-19 patients using only readily available variables.

### Clinical-GASS score and risk factors

The Clinical-GASS score is based on 11 variables easily available after the first visit at the Emergency Room, with the execution of both a venous and an arterial blood sample. The need for reliable and easy-to-use tools, able to predict the outcomes in COVID-19 inpatients is day by day more urgent because of the scarceness of human and financial resources, already under strain from the beginning of the pandemic.

The retrospective analyses from the first 499 patients hospitalized with COVID-19 to our hospitals in the territory of Ferrara, allowed us to notice a strong relationship between the Clinical-GASS score and mortality by COVID-19 (both in-hospital and 30-day rates); moreover, a slight relationship with the need for intensification of care was observed and confirmed only in the population with a Clinical-GASS score lower than 10 points; in the current study, this was true only for extreme values of the score (<5 or >10 points).

In our cohort of patients, the in-hospital mortality rate was 24.8%, while on the 30^th^ day of observation, it was 23.6%. Our findings showed how patients who underwent intensification of care, needing non-invasive mechanical ventilation or endotracheal intubation, were more often males, with a greater Clinical-GASS score (*p*<0.001) and worse respiratory performances (higher respiratory rates and lower PaO2/FiO2 ratios). As for laboratory abnormalities, those patients had higher serum inflammatory markers (CRP and ferritin), higher organ damage markers (isoamylase, ALT, LDH, HS TnI), and marked pro-coagulative status (higher serum D-Dimer).

Such findings are consistent with previous studies: male sex has been already linked to poor chances of survival from COVID-19 and with increased risk of admission to the Intensive Care Units (ICUs)[29] ^29^; similarly, PaO2/FiO2 ratio was already shown to be independently associated with worse COVID-19 outcomes[30] ^30^. Furthermore, recent studies showed how COVID-19 inpatients present commonly with laboratory abnormalities and the role of inflammatory, and organ-damage markers has been also consistently reported[31,32] ^31,32^. Our analyses also confirmed the role of age. Early Chinese studies showed how the elderly population was bound to encounter worse COVID-19 outcomes[33] ^33^, and such findings were later confirmed in studies with European[34,35] ^34,35^ and American patients[36] ^36^. Interestingly, the laboratory findings and respiratory performances reported in these studies were similar to those of our IoC group. Additionally, in accordance with previous studies that showed how low levels of blood pressure at hospital admission are often associated with worse COVID-19 outcomes[37] ^37^, we observed higher blood pressure (both systolic and diastolic) in the survivor group. Finally, the negative role of comorbidities (especially cardiovascular) and smoke[38] ^38^ is also well established[39,40] ^39,40^.

### Alternative clinical scores

Popular clinical scores of general applicability, such as the Charlson Comorbidity Score (CCI)[4] ^41^, the modified Elixhauser Index (mEI)[41] ^42^, the National Early Warning Score 2 (NEWS2)[42] ^43^ and the CURB models[8,9] ^8,9^ have been used during the current pandemic to predict COVID-19 outcomes with discrete success. However, they tend to underperform COVID-19-specific clinical scores[10,11]^10-12^.

The first COVID-19-specific scores were developed in China[43,44] ^44,45^, uring the first wave of the pandemic. Further scores for prediction of COVID outcomes were later developed all over the world[12,45,46] ^12,46,47^, and data from more heterogeneous datasets were collected in order to reduce potential selection bias stemming from sampling from a restricted population of subjects. A case in point is represented by the 4C score, developed by Knight et al.[12] ^12^: such a score has quickly become popular due to its vast derivation and validation cohorts and its ease of use. However, the predictive performance of the score (validation AUC = 0.77), appears to lag behind the one of SVM22-GASS introduced in this work (validation AUC = 0.87, accuracy = .88), and to be comparable with the one of the Clinical-GASS (validation AUC = 0.77, accuracy = 0.78). Similar arguments can be made with regard to the recently developed “Piacenza score” (validation AUC = 0.78, accuracy = 0.55).

Importantly, the SVM22-GASS bases its prediction on the analysis of clinical variables that can be quickly and inexpensively retrieved early on during hospital admission. This is a significant advantage over other popular multivariate methods that rely on the manual analysis of medical imaging results (e.g.,[11] ^11^), and yet achieve comparable predictive performance (validation AUC = 0.88). Like most machine-learning-based classifiers, the SVM22-GASS, the 4C and the Piacenza scores are purely data-driven. This makes them carry an intrinsic limitation represented by the fact that often it can be hard to give a “clinical” sense to the weight assigned to each feature considered in the development of the score. For this reason, in our first work, we decided to develop the GASS score, an informatic tool with the ability to accurately predict COVID-19 outcomes while keeping a clinical sense. The 11 features were chosen, in fact, based on both the strength of the associations with the outcomes and the clinical importance established for each of them in the current literature.

### Interpreting the SVM22-GASS classifier

The SVM22-GASS predicts the 30-day mortality outcome by nonlinearly processing 38 patient features. Our variable importance analysis isolated eight variables as the most important for the model: White Blood Cell Count (WBC), Lymphocyte Count (LYM), Brain Natriuretic Peptide (BNP), Creatine Phosphokinase (CPK), Lactate Dehydrogenase (LDH), Fibrinogen (FIBR), PaO2/FiO2 Ratio, and (PFR), and High-Sensitivity Troponin I (TnI). Interestingly, half of them, (namely, LYM, BNP, PFR, and TnI) are also part of the Clinical-GASS score, which confirms their strong information content and predictive power. Consistently, serum BNP has been recently found to be significantly elevated in critically ill COVID patients in a recent meta-analysis[47] ^48^; whether or not this peptide can help discriminate high-risk COVID-19 patients remains unclear and it merits further investigation. Similarly, high WBC values, high LDH values[15] ^15^, and low LYM values[15] ^15^, and have also been recently linked to high mortality risk. On the other hand, variables like age, sex, respiratory rate and serum D-Dimer (XDP) do not appear to influence significantly the predictions of the SVM22-GASS classifier. This finding might appear surprising, as their association with poor COVID-19 outcome is well established. For example, age is among the predominant risk factors for developing the severe form of the disease, due to the immuno-senescence and all the physiological modifications related to it[33] ^33^. Similarly, the serum levels of D-Dimer (XDP) were found to be strictly associated with COVID mortality[48] ^49^. Furthermore, the respiratory rate is a useful indicator of potential respiratory disfunctions. Nevertheless, the fact that the SVM22-GASS does not rely on such variables, does not mean that they provide no information about the mortality risk; it merely means that other variables, those that carry greater weight, are more informative and/or less noisy.

To validate the results of the variable importance analysis, we trained two additional classifiers: the SVM-TOP3 — which had only access to the three most important features White Blood Cell Count (WBC), Lymphocyte Count (LYM), Brain Natriuretic Peptide (BNP) — and the SVM-BOT3 — which had only access to the three least important features. Interestingly, we observed only a moderate performance decrease of the SVM-TOP3 with respect to the full SVM22-GASS model (AUC: 0.83 vs. 0.87, accuracy: 0.83 vs. 0.88). This suggests that SVM-TOP3 is also a competent predictor and can be used in those cases in which one does not have access to all of the 38 variables for a given patient.

### Potential Limitations

In this prospective cohort study, we used independent derivation and validation cohorts to develop and validate our COVID-19-specific classifiers and risk score. However, both cohorts were recruited from the same two hospitals in the Italian Province of Ferrara and included only participants of Caucasian ethnicity. This might lead to overestimating the performance of our methods. Additionally, the sample size we used, albeit comparable to that of many other related studies (e.g., see[46] ^47^), is still too small to allow to draw any final conclusion concerning the generalization ability of our approaches. To deal with these issues, future work will focus on undertaking a larger and multicentered study to validate our approaches on a larger and more heterogeneous cohort. Furthermore, in the current study we did not use information about medical treatments, as this was missing. This implies that we cannot determine whether and how drug prescriptions altered the prognostic trajectories of our patients.

## Conclusions

This work introduced two reliable classification approaches for the rapid assessment of mortality risk of COVID-19 patients. Importantly, both approaches rely on the automatic analysis of only routine clinical variables that can be readily acquired during hospital admission. The classifiers were developed and validated with independent derivation and validation cohorts and exhibited classification performance comparable to that of popular approaches that leverage information obtained with expensive medical imaging methods (e.g., chest radiography[11] – ^11^).

Our results prove that the classifiers have the potential to facilitate the triaging of incoming COVID-19 patients, thereby optimizing the allocation of hospital resources. Future work will further validate the proposed methods using larger and more heterogeneous validation cohorts, and will focus on designing suitable strategies to assess them in hospital settings.

## Data Availability

All data produced in the present study are available upon reasonable request to the authors

## Acknowledgements

Special thanks to the directors of the medical departments where our research took place: Giovanni Zuliani MD, PhD director of the Department of University Internal Medicine in the “Arcispedale S.Anna” in Cona (Fe), Roberto Manfredini MD, PhD, director of the Department of Clinical Medicine in the “Arcispedale S.Anna” in Cona (Fe) and Stefano Parini MD, director of the Department of Internal Medicine in the “Ospedale del Delta” in Lagosanto (Fe).

We would like to dedicate our sincere thanks to all the members of the medical staff that is still participating in the fight against the virus, to all the extremely skillful and valuable nursing staff members as well as our assistance personnel, every day working on the frontline.

Finally, the authors thank the International Max Planck Research School for Intelligent Systems (IMPRS-IS) for supporting AS.

