## Supplementary Table for "Rapid assessment of COVID-19 mortality risk with GASS classifiers"

|  | **0** | **+1** | **+2** | **+3** | **+4** |
| --- | --- | --- | --- | --- | --- |
| **Age (years)** | <45 | 45-60 | 61-74 | 75-89 | ≥90 |
| **Sex** | Female | Male |  |  |  |
| **RR (bpm)** | <20 | 20-30 | >30 |  |  |
| **PaO2/FiO2 ratio** | >300 | 200-300 | 100-199 | <100 |  |
| **Lymphocytes count (n/mmc)** | >1000 | 500-999 | <500 |  |  |
| **D-Dimer (mg/L FEU)** | <0.5 | 0.5-2.0 | >2.0 |  |  |
| **CKI** | Moderate | Severe |  |  |  |
| **eGFR (ml/min)** | >60 | 45-60 | 30-44 | <30 |  |
| **Troponin (ng/ml)** | <20 | 20-50 | >50 |  |  |
| **BNP (pg/ml)** | <100 | >100 |  |  |  |
| **CRP (mg/dl)** | 0-4.9 | 5.0-10.0 | >10.0 |  |  |

**GASS score items** – RR= Respiratory Rate; bpm= breaths per minute; CKI= Chronic Kidney Injury; eGFR= estimated Glomerular Filtration Rate; BNP= Brain Natriuretic Peptide; CRP= C Reactive Protein

|  | | **+1** | **+2** | | **+3** | **+6** |
| --- | --- | --- | --- | --- | --- | --- |
| **Coronary Artery Disease** | | * |  | |  |  |
| **Congestive Heart Failure** | | * |  | |  |  |
| **Chronic Pulmonary Disease** | | * |  | |  |  |
| **Peptic Ulcer Disease** | | * |  | |  |  |
| **Peripheral Vascular Disease** | | * |  | |  |  |
| **Mild Liver Disease** | | * |  | |  |  |
| **Cerebrovascular Disease** | | * |  | |  |  |
| **Connective Tissue Disease** | | * |  | |  |  |
| **Diabetes** | | * |  | |  |  |
| **Dementia** | | * |  | |  |  |
| **Hemiplegia** | |  | * | |  |  |
| **Moderate-to-Severe Renal Disease** |  |  | | * |  |  |
| **Diabetes with End-organ Damage** |  |  | | * |  |  |
| **Any Prior Tumour (within 5 yrs of Diagnosis)** |  |  | | * |  |  |
| **Leukaemia** |  |  | | * |  |  |
| **Lymphoma** |  |  | | * |  |  |
| **Moderate-to-Severe Liver Disease** |  |  | |  | * |  |
| **Metastatic Solid Tumour** |  |  | |  |  | * |
| **AIDS (not only HIV Positive)** |  |  | |  |  | * |

**Charlson Comorbidity Index items –** AIDS= Acquired ImmunoDeficiency Syndrome

| Variable | IoCp (N=76) | NIoCp (N=174) | *p* |
| --- | --- | --- | --- |
| Men | 50 (65.8) | 87 (50.0) | 0.021 |
| Women | 26 (34.2) | 87 (50.0) |  |
| Age <45 | 3 (3.9) | 25 (14.4) | 0.016 |
| Age 45-60 | 13 (17.1) | 36 (20.7) | 0.51 |
| Age 61-74 | 29 (38.2) | 33 (19.0) | 0.001 |
| Age 75-89 | 31 (40.8) | 65 (37.4) | 0.61 |
| Age ≥90 | 0 (0.0) | 15 (8.6) | 0.008 |
| RR <20 bpm | 11 (14.5) | 69 (39.7) | <0.001 |
| RR 20-30 bpm | 29 (38.2) | 60 (34.5) | 0.61 |
| RR >30 bpm | 19 (25.0) | 4 (2.3) | <0.001 |
| PaO2/fiO2 ratio >300 | 9 (11.8) | 77 (44.3) | <0.001 |
| PaO2/fiO2 ratio 200-300 | 18 (23.7) | 40 (23.0) | 0.81 |
| PaO2/fiO2 ratio 100-199 | 23 (30.3) | 8 (4.6) | <0.001 |
| PaO2/fiO2 ratio <100 | 11(14.5) | 3 (1.7) | <0.001 |
| Lymphocytes >1000/mmc | 32 (42.1) | 91 (52.3) | 0.11 |
| Lymphocytes 500-999/mmc | 34 (44.7) | 70 (40.2) | 0.58 |
| Lymphocytes <500/mmc | 10 (13.2) | 11 (6.3) | 0.08 |
| CRP <5 mg/dl | 31 (40.8) | 88 (50.6) | 0.16 |
| CRP 5-10 mg/dl | 12 (15.8) | 47 (27.0) | 0.06 |
| CRP >10 mg/dl | 32 (42.1) | 37 (21.3) | 0.001 |
| eGFR >60 ml/min | 54 (71.1) | 118 (67.8) | 0.55 |
| eGFR 45-60 ml/min | 12 (15.8) | 19 (10.9) | 0.27 |
| eGFR 30-44 ml/min | 5 (6.6) | 13 (7.5) | 0.81 |
| eGFR <30 ml/min | 4 (5.3) | 23 (13.2) | 0.06 |
| D-dimer <0.5 mg/L FEU | 8 (10.5) | 48 (27.6) | 0.002 |
| D-dimer 0.5-2 mg/L FEU | 47 (61.8) | 78 (44.8) | 0.035 |
| D-dimer >2 mg/L FEU | 21 (27.6) | 39 (22.4) | 0.51 |
| HS TnI <20 ng/ml | 42 (55.3) | 107 (61.5) | 0.49 |
| HS TnI 20-50 ng/ml | 13 (17.1) | 29 (16.7) | 0.85 |
| HS TnI >50 ng/ml | 14 (18.4) | 27 (15.5) | 0.50 |
| BNP <100 pg/ml | 41 (53.9) | 95 (54.6) | 0.94 |
| BNP ≥100 pg/ml | 27 (35.5) | 64 (36.8) | 0.94 |
| Clinical GASS <5 pts | 4 (5.3) | 49 (28.2) | <0.001 |
| Clinical GASS 5-10 pts | 42 (55.3) | 88 (50.6) | 0.50 |
| Clinical GASS >10 pts | 30 (39.5) | 37 (21.3) | 0.003 |

**GASS score items – differences between groups in terms of Intensification of Care.** IoCp = patients who needed Intensification of Care; NIoCp = patients who did not need Intensification of Care; RR = Respiratory Rate; CRP = C Reactive Protein; eGFR = estimated Glomerular Filtration Rate; HS TnI = High Sensitivity Troponin I; BNP = Brain Natriuretic Peptide; GASS = General Assessment of SARS-CoV-2 Severity

| Variable | Deceased (N=62) | Discharged (N=188) | *p* |
| --- | --- | --- | --- |
| Men | 31 (50.0) | 106 (56.4) | 0.38 |
| Women | 31 (50.0) | 82 (43.6) |  |
| Age <45 | 0 (0.0) | 28 (14.9) | 0.001 |
| Age 45-60 | 3 (4.8) | 46 (24.5) | 0.001 |
| Age 61-74 | 9 (14.5) | 53 (28.2) | 0.031 |
| Age 75-89 | 44 (71.0) | 52 (27.7) | <0.001 |
| Age ≥90 | 6 (9.7) | 9 (4.8) | 0.16 |
| RR <20 bpm | 14 (22.6) | 66 (35.1) | 0.06 |
| RR 20-30 bpm | 24 (38.7) | 65 (34.6) | 0.46 |
| RR >30 bpm | 9 (14.5) | 14 (7.4) | 0.08 |
| PaO2/fiO2 ratio >300 | 10 (16.1) | 76 (40.4) | 0.001 |
| PaO2/fiO2 ratio 200-300 | 17 (27.4) | 41 (21.8) | 0.12 |
| PaO2/fiO2 ratio 100-199 | 11 (17.7) | 20 (10.6) | 0.052 |
| PaO2/fiO2 ratio <100 | 4 (6.5) | 10 (5.3) | 0.55 |
| Lymphocytes >1000/mmc | 19 (30.6) | 104 (55.3) | 0.001 |
| Lymphocytes 500-999/mmc | 33 (53.2) | 71 (37.8) | 0.029 |
| Lymphocytes <500/mmc | 9 (14.5) | 12 (6.4) | 0.044 |
| CRP <5 mg/dl | 21 (33.9) | 98 (52.1) | 0.013 |
| CRP 5-10 mg/dl | 16 (25.8) | 43 (22.9) | 0.62 |
| CRP >10 mg/dl | 24 (38.7) | 45 (23.9) | 0.022 |
| eGFR >60 ml/min | 28 (45.2) | 144 (76.6) | <0.001 |
| eGFR 45-60 ml/min | 11 (17.7) | 20 (10.6) | 0.13 |
| eGFR 30-44 ml/min | 7 (11.3) | 11 (5.9) | 0.14 |
| eGFR <30 ml/min | 15 (24.2) | 12 (6.4) | <0.001 |
| D-dimer <0.5 mg/L FEU | 4 (6.5) | 52 (27.7) | 0.001 |
| D-dimer 0.5-2 mg/L FEU | 35 (56.5) | 90 (47.9) | 0.19 |
| D-dimer >2 mg/L FEU | 20 (32.3) | 40 (21.3) | 0.07 |
| HS TnI <20 ng/ml | 21 (33.9) | 128 (68.1) | <0.001 |
| HS TnI 20-50 ng/ml | 16 (25.8) | 26 (13.8) | 0.030 |
| HS TnI >50 ng/ml | 21 (33.9) | 20 (10.6) | <0.001 |
| BNP <100 pg/ml | 17 (27.4) | 119 (63.3) | <0.001 |
| BNP ≥100 pg/ml | 42 (67.7) | 49 (26.1) | <0.001 |
| GASS <5 pts | 2 (3.2) | 51 (27.1) | 0.53 |
| GASS 5-10 pts | 27 (43.5) | 103 (54.8) | 0.65 |
| GASS >10 pts | 33 (53.2) | 34 (18.1) | 0.82 |

**GASS score items – differences between groups in terms of in-hospital mortality.** RR = Respiratory Rate; CRP = C Reactive Protein; eGFR = estimated Glomerular Filtration Rate; HS TnI = High Sensitivity Troponin I; BNP = Brain Natriuretic Peptide; GASS = General Assessment of SARS-CoV-2 Severity

| Variable | 30-ddp (N=59) | 30-dsp (N=191) | *p* |
| --- | --- | --- | --- |
| Men | 28 (47.5) | 109 (57.1) | 0.20 |
| Women | 31 (52.5) | 82 (42.9) |  |
| Age <45 | 0 (0.0) | 28 (14.7) | 0.002 |
| Age 45-60 | 3 (5.1) | 46 (24.1) | 0.001 |
| Age 61-74 | 13 (22.0) | 49 (25.7) | 0.45 |
| Age 75-89 | 37 (62.7) | 59 (30.9) | <0.001 |
| Age ≥90 | 6 (10.2) | 9 (4.7) | 0.12 |
| RR <20 bpm | 16 (27.1) | 64 (33.5) | 0.42 |
| RR 20-30 bpm | 20 (33.9) | 69 (36.1) | 0.89 |
| RR >30 bpm | 8 (13.6) | 15 (7.9) | 0.15 |
| PaO2/fiO2 ratio >300 | 11 (18.6) | 75 (39.3) | 0.007 |
| PaO2/fiO2 ratio 200-300 | 15 (25.4) | 43 (22.5) | 0.36 |
| PaO2/fiO2 ratio 100-199 | 10 (16.9) | 21 (11.0) | 0.12 |
| PaO2/fiO2 ratio <100 | 5 (8.5) | 9 (4.7) | 0.19 |
| Lymphocytes >1000/mmc | 21 (35.6) | 102 (53.4) | 0.018 |
| Lymphocytes 500-999/mmc | 29 (49.2) | 75 (39.3) | 0.16 |
| Lymphocytes <500/mmc | 8 (13.6) | 13 (6.8) | 0.10 |
| CRP <5 mg/dl | 21 (35.6) | 98 (51.3) | 0.037 |
| CRP 5-10 mg/dl | 15 (25.4) | 44 (23.0) | 0.69 |
| CRP >10 mg/dl | 22 (37.3) | 47 (24.6) | 0.052 |
| eGFR >60 ml/min | 27 (45.8) | 145 (75.9) | <0.001 |
| eGFR 45-60 ml/min | 12 (20.3) | 19 (9.9) | 0.031 |
| eGFR 30-44 ml/min | 5 (8.5) | 13 (6.8) | 0.65 |
| eGFR <30 ml/min | 14 (23.7) | 13 (6.8) | <0.001 |
| D-dimer <0.5 mg/L FEU | 4 (6.8) | 52 (27.2) | 0.001 |
| D-dimer 0.5-2 mg/L FEU | 38 (64.4) | 87 (45.5) | 0.010 |
| D-dimer >2 mg/L FEU | 15 (25.4) | 45 (23.6) | 0.78 |
| HS TnI <20 ng/ml | 18 (30.5) | 131 (68.6) | <0.001 |
| HS TnI 20-50 ng/ml | 16 (27.1) | 26 (13.6) | 0.015 |
| HS TnI >50 ng/ml | 21 (35.6) | 20 (10.5) | <0.001 |
| BNP <100 pg/ml | 17 (28.8) | 119 (62.3) | <0.001 |
| BNP ≥100 pg/ml | 39 (66.1) | 52 (27.2) | <0.001 |
| GASS <5 pts | 2 (3.4) | 51 (26.7) | <0.001 |
| GASS 5-10 pts | 26 (44.1) | 104 (54.5) | 0.16 |
| GASS >10 pts | 31 (52.5) | 36 (18.8) | <0.001 |

**GASS score items – differences between groups in terms of 30-day mortality.** 30-ddp = patients deceased within 30 days; 30-dsp = patients survived after 30 days; RR = Respiratory Rate; CRP = C Reactive Protein; eGFR = estimated Glomerular Filtration Rate; HS TnI = High Sensitivity Troponin I; BNP = Brain Natriuretic Peptide; GASS = General Assessment of SARS-CoV-2 Severity
